# The Safety of Contemporary Planned Cancer Surgery During the COVID-19 Pandemic

**DOI:** 10.1101/2020.10.19.20207803

**Authors:** D Shackley, S Gray, S Penney, L Galligan-Dawson, F Howle, S Arya, J Hunter, F Andrews, C Wasson, M Luckas, B Ryan, J Eddleston, C Brookes, C Harrison

## Abstract

**Introduction:** The COVID-19 pandemic caused by the novel coronavirus SARS-CoV-2 has put an unprecedented burden on global healthcare, including detrimental implications for the volume and provision of surgical services. The aim of this audit was to assess if the planned surgical cancer care (both diagnostic for possible cancer, and treatment of known cancer) during this period of widespread community and hospital based COVID-19 infection resulted in patients acquiring symptomatic COVID-19 as a consequence of their surgical admission, and if so, what the impact on patients was.

**Methods:** A prospective audit of all patients undergoing elective cancer surgery in Greater Manchester operated on between 01/05/2020 and 31/06/2020 was undertaken after the introduction of specific peri operative COVID safety measures across Greater Manchester cancer surgical cells. The COVID related outcomes for all cancer patients operated on in Greater Manchester were recorded.

**Results:** Of the 1501 patients undergoing surgery, one (<0.1%) was diagnosed with COVID-19 in hospital within 14 days of surgery. This patient did not require admission to critical care due to post-operative COVID-19 diagnosis, and there was no associated mortality related to post-operative COVID-19 infection.

**Conclusion:** The use of peri operative COVID-19 infection prevention strategies has allowed for the safe continuation of elective cancer surgery during this pandemic in all surgical units, without significant additional COVID-19 related morbidity or mortality.

## Introduction

The COVID-19 pandemic caused by the novel coronavirus SARS-CoV-2 has put an unprecedented burden on global healthcare, including detrimental implications for the volume and provision of surgical services. Considerable surgical resources including staff, facilities and equipment (such as ventilators) were initially re-directed towards the preparation for, and the treatment of, symptomatic infected COVID-19 patients with the volumes of surgically-treated cases falling significantly across all specialities.

Early research highlighted significant concerns regarding the safety of surgery during the pandemic; Lei et al ^1^ published the results of a cohort of 34 patients undergoing elective surgery in January and February 2020 in Wuhan, China, who were unintentionally scheduled for elective surgery during the incubation period of COVID-19, all of whom developed symptomatic disease post-operatively. They reported a 44% critical care admission rate post operatively, with a 20% mortality rate. The ‘COVIDSurg collaborative group’ looked at 1128 patients in an international cohort who developed SARS-CoV-2 infection in the 7 days leading up to, or 30 days following surgery: They found a 23.8% mortality rate in patients diagnosed with COVID-19 in the peri-operative period. ^2^

A blunt strategy for the avoidance of this risk would be to delay surgery altogether. It was forecast that globally this would mean 28 million elective operations would be cancelled over the 12-week peak of the coronavirus pandemic, including over 2 million cancer operations. ^3^ The cancellation and delay of cancer surgery has devastating consequences for a health system, including unnecessary deaths. ^4,5^

These studies taken together highlighted the (1) morbidity and mortality associated with having COVID-19 infection around the time of surgery and (2) the scale and numbers of patients possibly affected who require cancer surgery. Further, they underlined the importance of pre-operative screening for COVID-19 in elective surgery, and to take all necessary steps to reduce the chance of acquiring COVID-19 in the peri-operative period in those where cancer surgery was performed.

In England, this led to the publication of (i) specific guidance for both the prioritisation of surgical procedures, and (ii) peri-operative measures/ guidance which should be taken to reduce the risk to patients of COVID-19 infection during the resumption of elective cancer surgery. ^6,7^ The clinical prioritisation tools/ guidance were used by local clinical committees based regionally or within Trusts across England to prioritise patients for cancer surgery based on urgency and clinical need. Greater Manchester was proactive in installing these steps which were largely in place from the end of April 2020.

The guidance described measures designed to reduce the risk of developing COVID-19 - related complications and harm and included requiring (1) a negative systematic COVID-19 screening test 72 hours prior to surgery, (2) the patient self-isolating for 14 days prior to surgery, (3) where possible, patients were managed in “COVID-Secure Green” facilities and cohorted into ward and theatre zones separate to acute admission areas where patients had not followed the same pre-admission protocol.

Elective cancer patients continued to be treated in Greater Manchester, United Kingdom throughout the pandemic in all Trusts following this protocol in the months of May and June 2020.

The Greater Manchester cancer system involves ten hospital Trusts serving a population of approximately 3 million across the North West of England. ^8^ At the time of writing, the North West has the highest COVID-19 infection rate over the course of the pandemic of any region in the United Kingdom at 699 cases per 100,000 population. ^9^ In the months of March to June 2020, the North West of England had the second highest age-standardised COVID-19 related mortality rate in the country, reporting 7,639 deaths. ^10^

The aim of this audit was to assess if the planned surgical cancer care (both diagnostic for possible cancer, and treatment of known cancer) during this period of widespread community and hospital based COVID-19 infection resulted in patients acquiring symptomatic COVID-19 as a consequence of their surgical admission, and if so, what the impact on patients was.

## Methods

Greater Manchester Surgical Principles were agreed by the Executive Medical Directors of all Trusts undertaking surgery across the city-region at the start of May 2020 following a review of available national guidance on appropriate safety measures for surgical patients.

These principles were formulated using the available published research, including guidance from NHS England ^6^ and the Royal College of Surgeons ^5^. This included the provision of:

- Systematic patient COVID-19 testing via a nasopharyngeal swab 72 hours prior to surgery for all patients - a negative test being required before admission
- Postponement of elective surgery for COVID-19 positive individuals with rescheduled surgery after they become COVID-negative on repeat swab testing
- The formation of “COVID-19 Secure” facilities with designated “Green” areas. Green areas being protected facilities used for the care of elective surgical patients who have been screened and are COVID-19 negative, separate from treatment areas where patients’ COVID-19 status is uncertain or positive (such as emergency care)
- Patients instructed to self-isolate in their own homes for 14 days prior to surgery
- Every effort made to use a dedicated elective pool of staff for these cases
- All staff instructed to inform their manager and ‘stay at home’ if they develop any symptoms that could be COVID-19 infection (staff testing protocol then initiated).
- An inpatient COVID-19 test was rapidly initiated in any surgical patient with possible symptoms and during the 2 month audit, a process was introduced where patients who remained in hospital for more than 7 days were routinely re-tested for SARS-CoV-2 infection

These procedures were implemented in each of the ten hospital trusts providing elective cancer surgery in May and June 2020. A prospective audit of all patients undergoing elective cancer surgery in Greater Manchester operated on between 01/05/2020 and 31/06/2020 was then undertaken as a standard measure to provide information and assurance to patients, healthcare teams and the wider public on the safety of contemporary surgery in a cancer cohort.

Inclusion criteria for the audit were defined as any patient undergoing elective cancer surgery, including diagnostic cancer surgery, during May and June 2020 with every effort being made to include all patients.

Exclusion criteria included emergency cases where admission was unplanned, non-cancer elective surgery and local anaesthetic cases. Each patient’s notes were accessed and audited for the following criteria:

a. Result of pre-operative COVID-19 nasopharyngeal screening swab within 72 hours of surgery
b. Post-operative hospital-based diagnosis of COVID-19 within 14 days of surgery via a positive nasopharyngeal swab
c. Admission to critical care due to diagnosis of COVID-19 post operatively
d. Mortality due to COVID-19 post operatively in the hospital setting

A root cause analysis (RCA) was performed on all patients diagnosed with COVID-19 in the post-operative period.

## Results

1512 (681 in May, 831 in June) elective cancer operations were scheduled in May and June 2020 across Greater Manchester. In the corresponding period for 2019, 2300 patients had cancer related surgery and so the volume of cancer surgery during this audit was 65% of the previous year’s surgical volume.

1505 patients had their COVID-19 swab results available prior to surgery with 7 patients proceeded to surgery without a COVID-19 swab result.

25 patients had a positive pre-operative COVID-19 swab and their surgery was re-arranged. Upon review 14 of these have had their re-scheduled surgery in May and June. Therefore 1501 elective cancer operations were performed in the two months analysed in this audit.

Table 1 sets out the types of major surgery performed in the 2 months in Greater Manchester

**Table 1.**
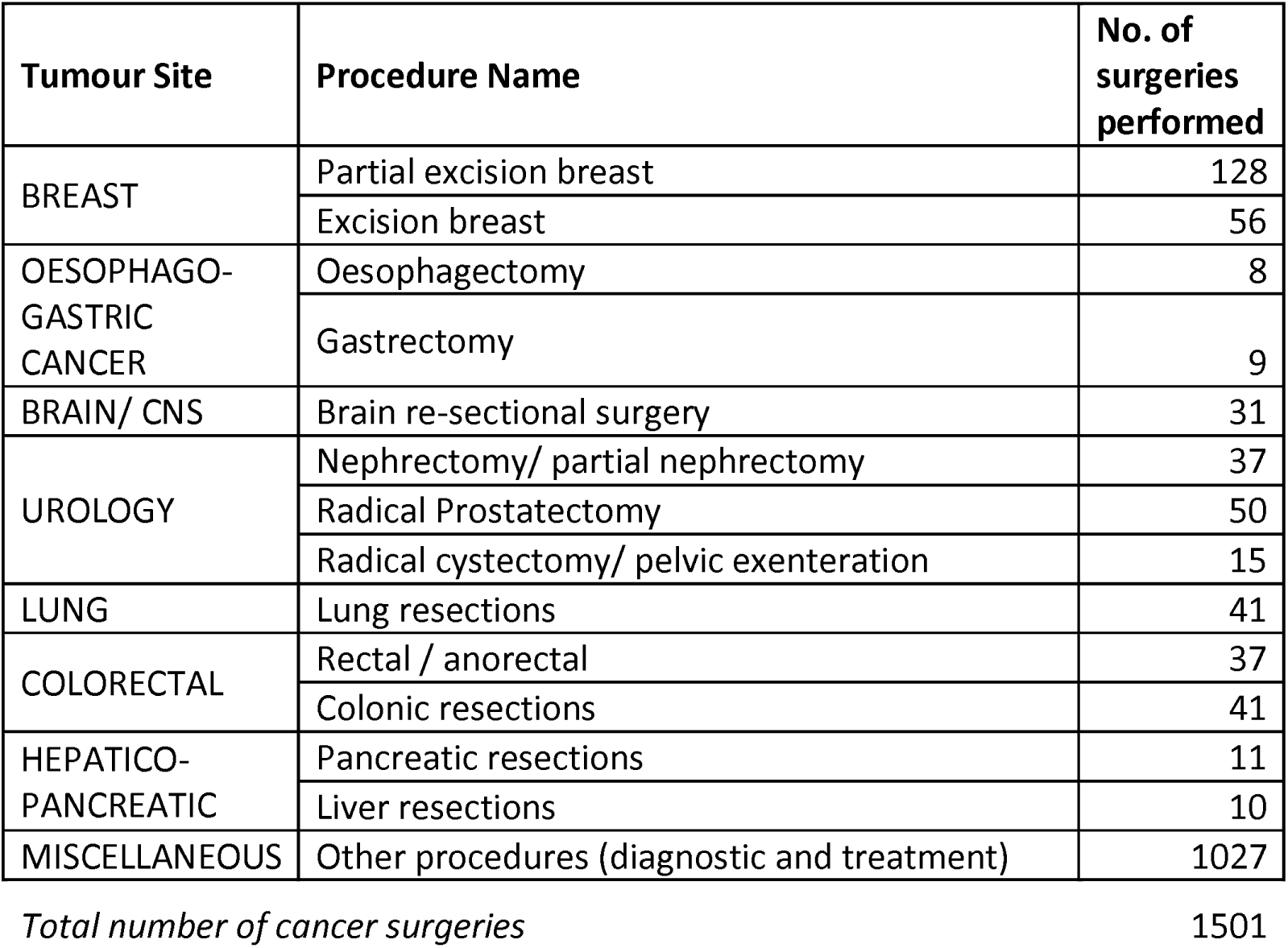
Cancer Surgery performed in Greater Manchester Cancer System in May & June 2020

Of the 1501 patients undergoing surgery, one (<0.1%) was diagnosed with COVID-19 in hospital within 14 days of surgery. This patient did not require admission to critical care due to post-operative COVID-19 diagnosis, and there was no associated mortality related to post-operative COVID-19 infection. None of the seven patients who underwent a procedure without a pre-operative screening COVID-19 swab developed known COVID-19 (as defined by a positive swab whist an in-patient). Table 2 sets out a summary of the key results.

**Table 2:** Summary of Key Results

| Number of Included patients | Number of patients with positive pre-operative COVID-19 swab | Number of patients preceding to surgery in May or June 2020 (including patients who were rescheduled for a later date due to initial positive pre-op swab) | Number of patients with post-operative inpatient positive COVID-19 swab within 14 days surgery | Number of patients with post-operative critical care admission or death due to COVID-19 infection |
| --- | --- | --- | --- | --- |
| 1512 | 25 | 1501 | 1 | 0 |

The single patient who acquired COVID-19 post operatively underwent a root cause analysis. This patient had a negative pre-operative screening swab and had a positive COVID-19 swab day 6 post laparoscopic colectomy (this test being scheduled for all inpatients remaining in at 5-7 days after admission in that trust). The patient was asymptomatic and measures were taken to discharge her on her expected date of discharge with self isolation advice. She remained asymptomatic and did not re-present to secondary care after discharge and was asymptomatic on telephone follow up.

Of the 25 patients who were cancelled due to a positive preoperative swab, follow up information is available for 22, and 19 have since had surgery with a median delay of 18 days (longest 50 days) with 2 patients removed from the waiting list due to complicating medical (non COVID-19) issues, and 1 removed due to patient choice to not have surgery currently and to be reviewed in September.

## Discussion

This study is the largest contemporary analysis of COVID-19 related outcomes for patients undergoing elective cancer surgery during this pandemic to date. It is the first of its kind to look at outcomes following the widespread introduction of COVID-19 prevention strategies peri-operatively.

The audit offers considerable assurance to healthcare teams, patients and the broader public that contemporary cancer surgery with the COVID-19 precautions outlined (principally 14 days self-isolation and a negative preoperative swab result within 72 hours of surgery) is ‘COVID-safe’.

Of the 1501 cancer patients who underwent cancer surgery, only 1 (<0.1%) acquired a proven post-operative in-patient SARS-CoV-2 diagnosis, and this patient was asymptomatic and picked up by a routine post-operative COVID-19 swab. This patient self-isolated on discharge for 14 days and on later review confirmed they had experienced no COVID-19 symptoms.

Twenty five patients (25/1501; 1.7%) were found to have positive swabs pre-operatively and their surgery was therefore delayed though 14 of these still had surgery within the audit period of May and June 2020.

Seven patients (0.5%) did not have a swab pre-operatively but went forward for surgery regardless, and none of these seven acquired COVID-19 symptoms in the post-operative period. These patients were from the start of the cohort when processes were being established. Robust processes are now in place in Greater Manchester to stop elective patients from receiving surgery without a negative COVID-19 swab within 72 hours of planned surgery.

The widely reported retrospective study published early in the pandemic by Lie et al ^1^ gave grave concern to the surgical community that operations may exacerbate very significant morbidity and mortality in COVID-19 infected individuals. It is widely known that immune system function and inflammatory response is important in the disease progression of this viral infection. ^11^ Surgery and anaesthesia have long been associated with impaired immunity ^12,13^ and a systemic inflammatory response. ^14^

This study confirms the effectiveness of Greater Manchester’s approach in adopting a set of clinically agreed bio-safety measures which has facilitated cancer surgery’s continuation throughout the crisis.

Of note is that all hospitals undertaking cancer surgery in the pre-COVID-19 era continued to do so following the UK national ‘lock-down’ and related NHS response of March 2020, though in reduced numbers.

The authors hope that this study can be used as evidence to encourage a broader resumption of planned elective surgery (cancer and non-cancer).

The limitations of this study design include potentially not identifying cases of asymptomatic COVID-19 positive patients post operatively or those who, after discharge, developed symptomatic COVID that did not require hospital admission. This is because all post operative patients were not routinely re-tested for COVID-19 unless symptoms or clinical deterioration occurred in a hospital setting within 14 days of surgery, or they remained in hospital for > 7 days (when routine testing was performed). An additional limitation is the unknown compliance of individuals with the 14 day pre-operative self-isolation protocol. Additionally, the bio-safety measures of green areas were adopted as ‘principles’ with every effort taken to avoid patient or staff–cross contamination but this could not be guaranteed in all settings particularly in the earlier phase of the audit period.

Although these concerns are valid, they had no detrimental effect on the COVID-19 related outcomes in our cohort of patients which remain favourable.

The strategies utilised for the peri operative screening and prevention of COVID-19 infection in Greater Manchester are replicable. However, it is important to bear in mind that peri operative patient self-isolation, designated COVID-secure facilities and systematic pre-operative COVID-19 screening requires significant resources and infrastructure. These resources include the economic loss of a patient adhering to 14 days of self-isolation, and compliance with this measure is likely to be variable based on an individual’s circumstance and the social support available in their nation. There are notable flaws in the accuracy of nasopharyngeal swabs for screening for COVID-19 infection and incubation ^12^, however these had little impact on discernible post-operative COVID-19 infections in our cohort of patients.

The COVID-19 pandemic has demanded changes to be made to the previous standard care for cancer patients across the world. This study supports the assertion that urgent cancer surgery can be safely performed during a COVID-19 pandemic provided certain safety precautions are in place.

## Conclusion

The use of peri operative COVID-19 infection prevention strategies has allowed for the safe continuation of elective cancer surgery during this pandemic in all surgical units, without significant additional COVID-19 related morbidity or mortality. This study can be used as part of individual patient risk discussions in relation to planned cancer surgery.

## Data Availability

All data available to public

